# Design of the Play & Grow Cohort: A prospective study of parent-child mealtime interactions

**DOI:** 10.1101/2020.06.11.20128637

**Authors:** Andria Parrott, Bharahi J. Zvara, Sarah A. Keim, Rebecca Andridge, Sarah E. Anderson

## Abstract

Early childhood is a critical period of life when nutritional choice and mealtime routines develop and obesogenic tendencies may be established. There is a need for greater understanding of toddlers and how parent-child interactions over time and across contexts relate to development of obesity. The purpose of this protocol paper is to describe the recruitment strategy, protocol, and characteristics of participants in the Play & Grow cohort. Between December 2017 and May 2019, three-hundred caregivers and their 16- to 19-month-old toddler (57% male) were recruited from records of the major pediatric provider in Central Ohio and enrolled in an ongoing 2-year prospective study. The cohort is diverse in race and ethnicity, parent education and age, household income, and family composition. Thirty-seven percent of children were born preterm (<37 weeks’ gestation). The first visit, completed by 299 families at mean (SD)18.2 (0.7) months child calendar age, included video-recorded parent-child interaction during play, anthropometric measurements, and caregiver questionnaire. Six months later, a home visit completed by 284 families included observation and video-recording of parent-child interaction in mealtime and non-mealtime contexts. This diverse cohort is being followed prospectively through ongoing home and laboratory visits at child ages 36 and 42 months. Growth trajectories of children will be analyzed in relation to self-regulation and quality of parent-child interaction.

## 1. INTRODUCTION

When children are young, families establish routines, set expectations, and develop patterns of interaction that shape future health behaviors and the home environment (Fiese & Bost, 2016). How parents feed children (feeding practices) can influence children’s food consumption and preferences, and are related to children’s weight gain and adiposity, although whether the child’s weight is the cause or consequence of the parent’s feeding practices is unclear (Spill et al., 2019). Premature birth has significant and lasting impacts on parenting and the parent-child relationship (Treyvaud, 2014). Preterm infants are smaller than their term peers for a given calendar age, and some struggle during the first few years of life to catch up to the growth of their peers born at term (Hack et al., 2003). Growth faltering is associated with poor outcomes, so clinical care for children born preterm often focuses on promoting growth (Franz et al., 2009). However, by school-age most children born preterm do catch up to be of similar size as children born at term (Hack et al., 2003). Despite continued monitoring of growth as part of clinical care for children born preterm, little research attention has been focused on preventing excess weight, an under-recognized consequence of growth-promoting behaviors that continue after adequate growth is achieved. Empirical research to assess the impact of preterm birth on parent feeding practices beyond infancy is also limited.

Childhood obesity is an important public health problem that has resisted easy solutions despite substantial efforts (Brown et al., 2019; Hales, Fryar, Carroll, Freedman, & Ogden, 2018). Children born preterm (<37 weeks’ gestation) have risks for obesity that are similar to those of children born full term (Casey et al., 2012; Cunningham, Kramer, & Narayan, 2014), and higher weight gain throughout the first year of life is linked to an increased risk for obesity in term (Geserick et al., 2018; Zheng et al., 2018) and preterm children (Belfort, Gillman, Buka, Casey, & McCormick, 2013). Obesity prevention efforts tailored to young children and their families hold promise (Lumeng, Taveras, Birch, & Yanovski, 2015), but there is a need for longitudinal research to understand the complex, reciprocal interactions through which parents create environments that shape children’s development and obesity risk (Baidal et al., 2016). In particular, toddlers are understudied relative to infants and school-age children. This paper describes the design of the Play & Grow cohort, a longitudinal study of diverse U.S. families with toddlers born both preterm and at term.

The emotional context of parent-child interactions and how caregiver responses to infant’s cues inform children’s recognition and interpretation of hunger has long been linked to obesity (Birch, Marlin, Kramer, & Peyer, 1981; Bruch, 1973; Fiese & Bost, 2016; Saltzman, Fiese, Bost, & McBride, 2018). Epidemiologic evidence suggests that risk for obesity is increased for young children who experience poor-quality emotional relationships with their parents, and these associations are not fully explained by differences in family socioeconomic position (Anderson, Gooze, Lemeshow, & Whitaker, 2012; Wendland et al., 2014; Wu, Dixon, Dalton, Tudiver, & Liu, 2011).

Decades of research by developmental scientists demonstrate that positive parent-child relationships support children’s development of self-regulation (Kochanska, Coy, & Murray, 2001). Prospective studies have suggested that children with lower self-regulation are more likely to be obese later in life (Francis & Susman, 2009; Graziano, Kelleher, Calkins, Keane, & Brien, 2013; Schlam, Wilson, Shoda, Mischel, & Ayduk, 2013), and children born preterm are at higher risk for deficits in self-regulation (Clark, Woodward, Horwood, & Moor, 2008). Difficulties with self-regulation could explain how poor-quality parent-child interactions increase obesity risk, but this has not been established, and which aspects of self-regulation are involved is only beginning to be explored (Anderson & Keim, 2016; Fiese & Bost, 2016; Hughes, Power, O’Connor, & Fisher, 2015). Further, parenting and child self-regulation may interact as predictors of children’s risk for obesity (Moding, Augustine, & Stifter, 2019). Research in diverse cohorts is needed to ensure that potential heterogeneity in associations guide development of theory (Bergmeier et al., 2020; Saltzman et al., 2018). Parent-child interactions are complex, and childhood obesity prevention efforts are strengthened by the increasing use of observational protocols to assess parent-child interactions broadly and across settings.

The extent to which parent-child interactions in the context of eating or mealtimes differ from how parents interact with their children in non-food settings has received surprisingly little research attention. An exception is Birch et al.’s cross-sectional study of 21 mothers and their preschool-aged child observed in a laboratory setting during lunch and while completing a puzzle task (1981). They report associations with child adiposity relative to patterns of mother-child interaction and conclude that fatter children experienced less responsive interactions in each context (i.e., eating or play) (Birch et al., 1981). In a recently published study of a cohort of infants at high risk due to maternal substance use, Kong and colleagues reported that warm and positive interactions between mothers and infants during play were associated with children’s lower BMI trajectories into elementary school, but quality of mother-infant interactions assessed during feeding were not associated with children’s BMI trajectories (Kong, Eiden, & Paluch, 2019). The quality of parent-child interaction in relation to child outcomes, such as obesity, has typically been assessed by coding parent and child behavior as observed during a semi-structured play task. Yet, it is plausible that parent-child interactions differ between mealtime and playtime contexts. Observations of diverse families over time with consistent measurement of parent-child interactions across contexts can inform the development of obesity prevention strategies targeting young children.

## 2. METHODS

### 2.1 Purpose and Setting

Play & Grow is a prospective cohort study of 300 caregiver-child dyads residing in Central Ohio in the United States. The primary goal of this NIH-funded study is to determine how the quality of parent-child interactions observed in mealtime and play settings in the home and laboratory impacts changes in weight and risk for obesity as children get older and identify the aspects of child self-regulation that are involved. The project is innovative in utilizing objective coding of observed parent-child interactions in mealtime and non-mealtime contexts over time and with children of all gestational ages at birth to better understand risk and protective factors for obesity and their interrelationships with self-regulation. This report describes the objectives of the study, recruitment of the cohort, protocol, and initial findings from the first 2 study visit time points when children were toddlers (18 months and 24 months of age). Hypotheses were established before data were collected, and data collection for this toddler phase of the study was completed in December 2019. The preschool phase of the study is ongoing.

### 2.2. Eligibility and Recruitment

Recruitment for the Play & Grow Cohort began in November of 2017 with approval from Nationwide Children’s Hospital’s Institutional Review Board (IRB16-00826). Families were recruited from two source populations using patient records maintained by Nationwide Children’s Hospital (NCH, Columbus, Ohio, USA). NCH is the only provider of subspecialty and emergency pediatric care in the region, and the major provider of pediatric primary care. NCH electronic medical records identified children whose calendar age was ≥16.0 and <17.0 months, and who had visited, at any point in their lives, an NCH Urgent Care Center, or who had been a patient in an NCH-affiliated Neonatal Intensive Care Unit (NICU) upon their preterm birth or been referred to the Neonatology Clinic for follow-up after a preterm birth. These two source populations (urgent care and NICU/Neonatology Clinic) were used to ensure sociodemographic diversity of participants across gestational ages.

#### Exclusion and Inclusion Criteria

Participating children were required to be born a singleton, be 18 ± 2 months chronological age at study enrollment, have their gestational age available in their medical record or reported by their caregiver if not available in the medical record, live within 15 miles of the NCH Main Campus in downtown Columbus with no family plans to move beyond that radius in the next 2 years; the child had to be able to communicate, self-feed, and move around the room during play. Additionally, the caregiver needed to be the child’s legal guardian, speak English with the child, and participate in the child’s meals on a regular basis. If two caregivers met these criteria, the primary caregiver was self-nominated and remained in that role across all visits. Participants were excluded if any of the following criteria were identified: child deafness, child blindness, a caregiver or child food allergy, the child’s recorded gestational age exceeded 42 weeks, or the child was tube-fed or a patient for a clinical feeding disorder. Additionally, children born at term who had been patients in the NICU were excluded. The rationale for this exclusion was the likelihood of severe clinical conditions associated with NICU admission for term neonates. Children whose medical record suggested they met inclusion and exclusion criteria were screened by study staff, and eligibility was confirmed with caregivers prior to enrollment. However, after recruitment we determined that four children who were born at term and had short stays (<7 days) at an NCH NICU as infants and had enrolled in the study. We decided to retain these children in the cohort because a review of their medical histories indicated that their stays in the NICU were not attributed to a severe health condition and their overall development reflected that of a healthy, typically developing child.

#### Recruitment Process

Recruitment took place between November 2017 and May 2019. The NCH database was queried monthly to generate a list of 100 to 150 children to attempt for recruitment; this included all age-eligible children whose gestational age was <35 completed weeks’ (approximately 30-40 children/month) and a random sample of children with unknown or later gestational ages at birth. In total, 2670 children were identified for potential recruitment (Figure 1). Of these, 671 children were not invited to participate because research staff identified an exclusion in their medical record prior to contact (n=294), or more children were eligible during a particular month than could be accommodated (n=377).

**Figure 1:**
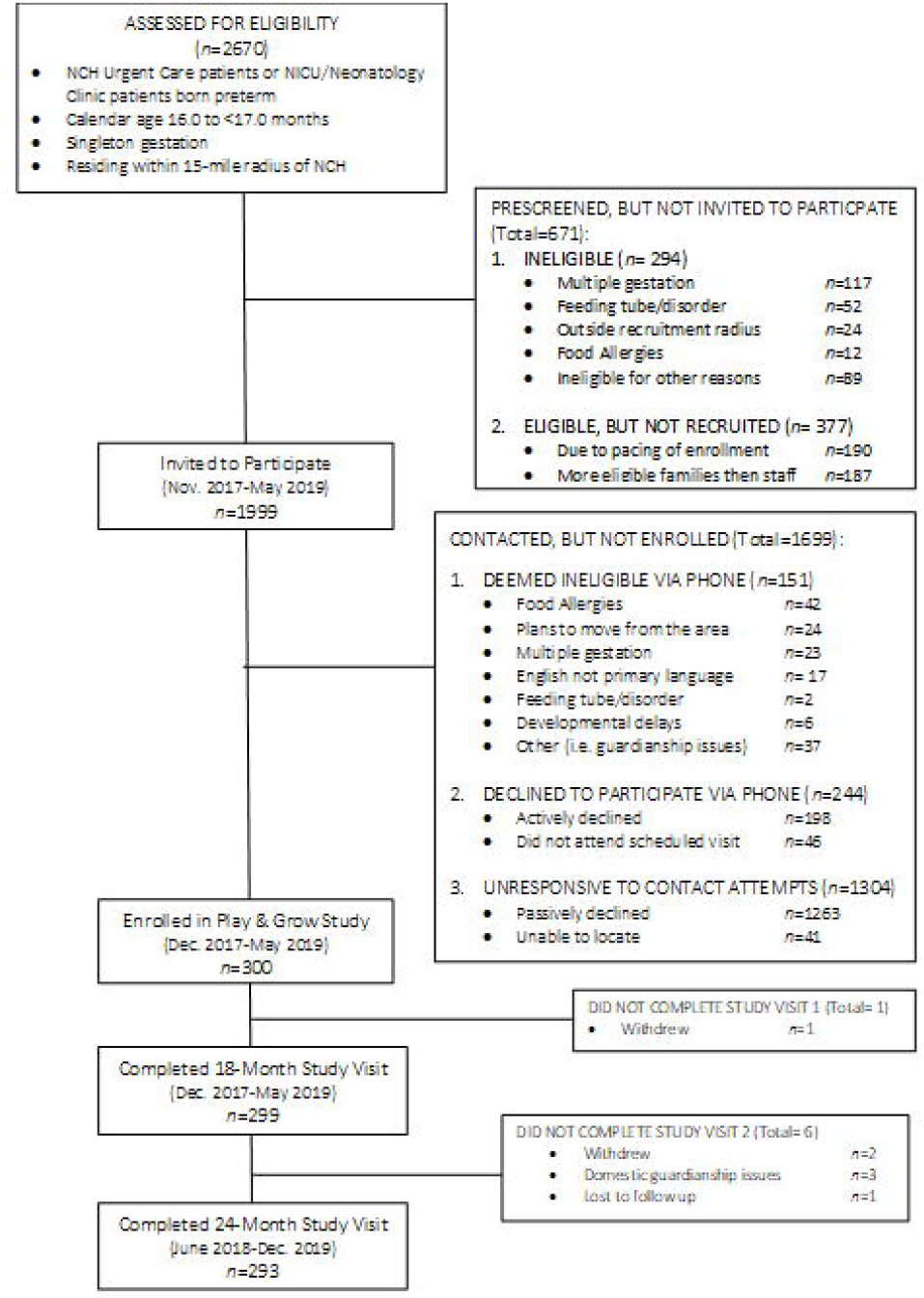
Play & Grow Study Recruitment and Flow Diagram.

Of the 1999 children and their caregivers invited to participate, 300 enrolled. To initiate recruitment, study staff identified the child’s legal guardian and contact information in the medical record. Recruitment began with a letter and a phone call one week later to gauge interest and further assess eligibility. If not reached during an initial call, contact attempts were repeated by phone, email or text message at varying times of the week and day. Eligible and interested families were scheduled to complete the informed consent process and enroll at the initial study visit. In total, 151 children and caregivers were deemed ineligible during a phone conversation with the caregiver, 198 caregivers actively declined to participate, and 46 scheduled a visit but did not attend. Recruitment efforts continued with each family until the child reached 20-months of age. In total, 1304 caregivers and children were unresponsive to contact attempts: 1263 of them passively declined by not responding, and 41 were not locatable with the available contact information (Figure 1). Recruitment was monitored by child sex, race and ethnicity to balance sociodemographic characteristics relative to children’s gestational age. Our goal was to enroll a diverse cohort that included children of all gestational ages and overrepresented children who were born very preterm (<32 completed weeks).

### 2.3 Data Collection

The toddler phase of the study included 2 visits separated by 6 months. The first visit coincided with enrollment and took place at the NCH Center for Biobehavioral Health when children were 16- to 19-months’ calendar age. The second study visit occurred in the participant’s home when the child was approximately 24-months old.

#### Protocol for First Visit

The primary caregiver and child attended the intial study visit at the NCH observational laboratory, gave written informed consent to participate in this 2-year longitudinal study consisting of 4 study visits (2 in the laboratory and 2 in their home), each involving various video-recorded tasks. Caregivers completed a 45-minute self-administered questionnaire on a tablet computer or on paper. Questions covered infant feeding practices, children’s daily routines, home environment, development, caregiver health and relationships, and household characteristics including food security. Study staff remained in the room to entertain the child and were available to answer caregiver questions.

Other components of the visit required 45 minutes and included anthropometric measurement of caregivers and toddlers (described in a later section) and a video-recording of parent-child interaction during a semi-structured play protocol (NICHD Early Child Care Research Network, 1999). Following standardized procedures, the caregiver and child were invited to sit on a floor mat, and a staff member presented a set of developmentally-appropriate toys (Fisher-Price® Little People Lil’ Movers™ Airplane, VTech® Busy Learners Activity Cube, and Sassy® Block Set, Zoomin’ Train). Caregivers were asked “*to play as you would at home if you had some free time*,” and were instructed to try to keep themselves and the child oriented with their faces and hands visible to the camera. Staff were absent during the parent-child interaction but remotely monitored the room for the 10-minute task duration. Videos were uploaded to a secure server. Recordings were observed and coded by independent teams of coders supervised and trained by an expert coder (BJZ). Coding teams were blinded to all other aspects of the family’s status and data and to our specific hypotheses. Interactions were coded (using a 7-point scale with 1= very uncharacteristic and 7= very characteristic) for the following global dimensions of parenting behavior: sensitivity, detachment, intrusive control, stimulation, warm positive regard and harsh negative regard, as well as overall mutuality of the interaction. This coding scheme is well-validated and has been used for mothers and fathers at varying levels of education and income as well as across ethnicities (Zvara et al., 2014). The visit concluded with a series of administrative tasks which included scheduling the home visit, thanking the caregiver with a $50 gift-card, book and study-branded blanket for the child, and facilitating transportation (i.e., parking validation or taxi) (Figure 2).

**Figure 2:**
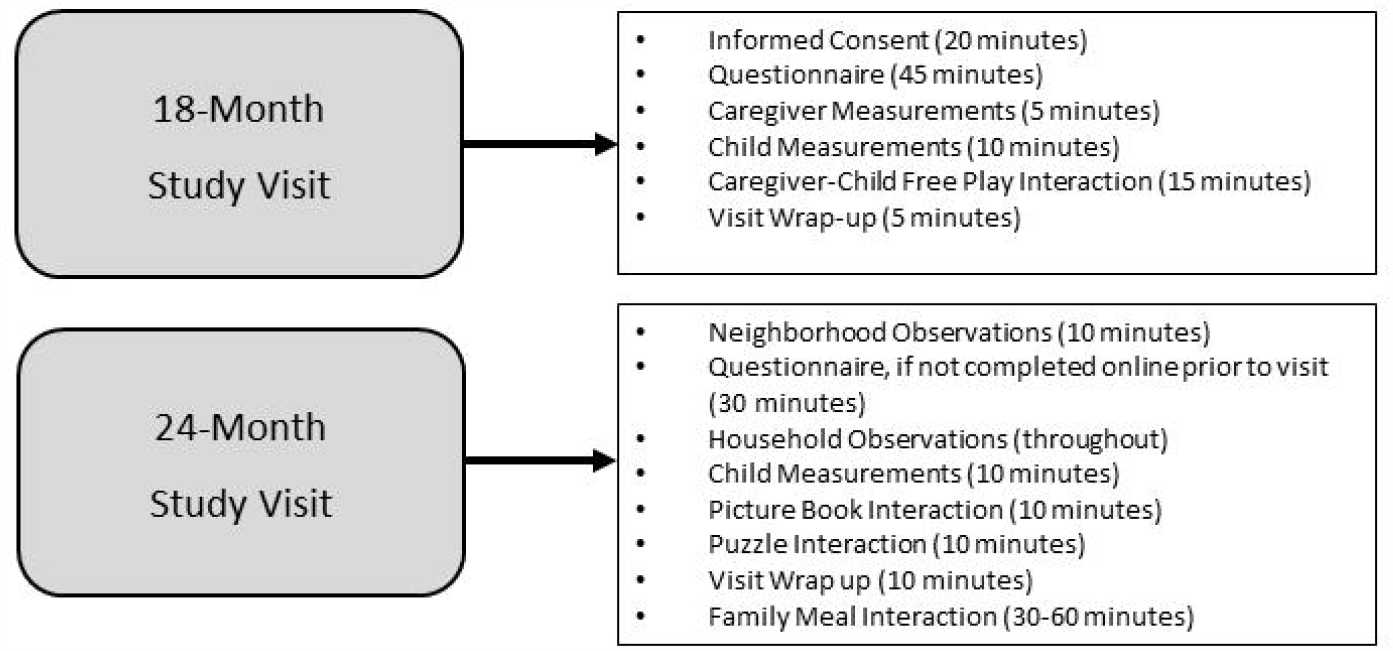
Activities at 18 & 24-Month Study Visits.

#### Anthropometric Measurements

Research staff received standardized training in adult and pediatric anthropometric measurement (Centers for Disease Control and Prevention, 2017), and were required to demonstrate accuracy and reliability in measurement of adult height and weight, and child weight, recumbent length, and standing height before they were certified to measure participants. Absolute technical error of measurement (TEM) was calculated for each trainee based on a minimum of 10 measurements (Perini, Oliveira, Ornellas, & Oliveira, 2005), and if staff did not obtain acceptable levels of accuracy and precision, they repeated trainings and measurements until they demonstrated proficiency.

#### Caregiver height and weight

The height and weight of the primary caregiver was measured at study visit 1. Height was measured to the nearest 0.1 cm using a Seca 284 stadiometer and weight was measured to the nearest 0.05 kg using a Seca 874 scale. Caregivers were dressed in light clothing without shoes. Equipment was calibrated prior to measurement. Height and weight measurements were taken three times according to a standardized procedure (Centers for Disease Control and Prevention, 2017). If the primary caregiver was pregnant, height was measured, and weight measured at a subsequent visit. In addition, when the primary caregiver was not the biological mother (n=20), we sought to measure the height and weight of the biological mother as well as the primary caregiver.

#### Child weight and length

The child was measured without shoes and in a clean diaper. Recumbent length was assessed to the nearest 0.1 cm using a calibrated Seca 416 infantometer. Research staff and the caregiver assisted with positioning the child, and 3 repeated length measurements were recorded. Child weight was taken in triplicate to the nearest 0.05 kg with the same Seca 874 scale used with caregivers.

#### Protocol for Second Visit

The second study visit occurred in the family’s home for approximately 90 minutes (Figure 2) and was scheduled for ±1 week surrounding the child’s second birthday. However, to maximize retention, caregivers were accommodated in the timing of the visit and we rescheduled visits until the child was 30-months old. The study provided food for a family meal if desired. Caregivers selected items from a Subway® Restaurants menu and staff brought the order to the home visit. However, 13% of families chose to provide their own food. Trained staff members completed the home visits in teams of two.

Caregivers were invited to complete a 30-minute self-administered questionnaire prior to the visit which assessed child and family routines, feeding strategies, neighborhood safety, and caregiver health behaviors as well as multiple aspects of parent physical, social, and emotional health and well-being.

Prior to and during the home visit, staff conducted a qualitative and quantitative assessment of neighborhood and household conditions. The methodology was adapted from existing instruments focused on neighborhood and home environments (Brownson et al., 2004; Caldwell & Bradley, 1984; Matheny Jr, Wachs, Ludwig, & Phillips, 1995; Ross & Mirowsky, 1999), and included observations of housing quality and maintenance, noise, safety, and amenities. The neighborhood was assessed for approximately 10 minutes prior to the start of the visit, and the home environment was observed throughout the visit (Figure 2).

Children’s standing height and weight were measured in triplicate using a portable Seca 213 Stadiometer and a Seca 874 scale. Each instrument was calibrated, and staff were trained to place them on a flat, level surface. The child was dressed in light clothing without shoes. Parent-child interaction was video-recorded in two 10-minute sessions; the first used a wordless picture book, and the second used three puzzles that ranged in difficulty. We concluded the visit by video-recording the child’s typical family meal (dinner for 53% and lunch for 47% of families). Video-recording lasted for 25-minutes or until the caregiver indicated that the meal was finished. Video recordings of all parent-child interactions (book, puzzles, meal) were uploaded to a secure server for observational coding.

### 2.4. Analytic Approach

The analytic plan for this study was pre-specified and all data-driven analyses were identified and discussed by the investigator team. We calculated body mass index (BMI) as weight (kg) divided by the square of height (m). Triplicate height and weight measurements were averaged, or if two measurements were identical, that value was used. We used BMI to categorize caregiver weight status: underweight (BMI <18.5 kg/m^2^), healthy weight (BMI ≥ 18.5 and <25 kg/m^2^), overweight (BMI ≥25 and <30 kg/m^2^), and obesity (BMI ≥30 kg/m^2^). Children’s weight status was defined relative to the World Health Organization (WHO) Child Growth Standards (WHO Multicentre Growth Reference Study Group, 2006). Sex-specific BMI-for-age z-scores were calculated using anthropometric measurements from each visit. Children’s age at each visit was calculated using date of birth, and if children were born before 37 completed weeks’ gestation, age was adjusted for prematurity. The WHO SAS macro was used to calculate BMI z-scores (World Health Organization, 2019).

We conducted descriptive analyses and present sociodemographic characteristics of children and caregivers. We examined the distribution of gestational age at birth and assessed differences in child and caregiver characteristics with chi-square tests and t-tests comparing children born preterm (<37 completed weeks’ gestation) to children born at term (≥37 completed weeks’ gestation). Analyses were conducted in SAS version 9.4 (SAS Institute, Cary, NC), and we used an alpha level of <0.05 to denote statistical significance.

## 3. RESULTS

### 3.1 Sociodemographic Characteristics

The 300 children (57% male) in the Play & Grow Cohort were born between June 2016 and December 2017 and were enrolled in the study at a mean (SD) [IQR] calendar age of 18.2 (0.70) [0.85] months. Children were born at gestational ages ranging 23 to 41 weeks’ completed gestation (Figure 3). The proportion of children born preterm (<37 weeks) was 37% (n=112) and this included 48 children born extremely or very preterm (<32 weeks). The primary caregiver was typically the biological mother (93%). The cohort includes diversity in child and caregiver race and ethnicity with a majority of children identified by their caregivers as having minority race or ethnicity (Table 1). Caregiver education ranged from high school degree or less (23%) to graduate degree (20%). Most caregivers (76%) were married or living with a partner. Household income varied widely; 26% had annual household incomes below $20,000 and 25% had incomes above $90,000. Household food security was low or very low for 17% of participants (Table 1).

**Table 1:** Child, Caregiver and Household Characteristics.

| **Child Characteristics** | **N (%)** |
| --- | --- |
| Gestational age at birth |  |
| 37-41 completed weeks' (term) | 188 (63%) |
| <37 completed weeks' (preterm) | 112 (37%) |
| Sex |  |
| Male | 170 (57%) |
| Female | 130 (43%) |
| Race/ethnicity |  |
| Non-Hispanic White | 137 (46%) |
| Non-Hispanic Black | 105 (35%) |
| Non-Hispanic other (includes multiple races) | 35 (12%) |
| Hispanic | 23 (8%) |
| **Caregiver Characteristics** | **N (%)** |
| Relationship to child |  |
| Biological mother | 280 (93%) |
| Biological father | 15 (5%) |
| Other<sup>a</sup> | 5 (2%) |
| Age (years) at enrollment |  |
| 18 to <21 | 8 (3%) |
| 21 to <25 | 46 (15%) |
| 25 to <30 | 62 (21%) |
| 30 to <35 | 98 (33%) |
| 35 to <40 | 62 (21%) |
| 40 or older | 23 (8%) |
| Race/ethnicity |  |
| Non-Hispanic White | 158 (53%) |
| Non-Hispanic Black | 111 (37%) |
| Non-Hispanic other (includes multiple races) | 18 (6%) |
| Hispanic | 13 (4%) |
| Marital status |  |
| Married | 162 (55%) |
| Living with partner | 62 (21%) |
| Single/never married | 58 (20%) |
| Other<sup>b</sup> | 15 (5%) |
| Education level |  |
| High school or less | 70 (23%) |
| Some college or Associate's degree | 103 (34%) |
| Bachelor's degree | 67 (22%) |
| Graduate degree | 59 (20%) |

| **Household Characteristics** | **N (%)** |
| --- | --- |
| Annual Household Income |  |
| <\$20 thousand | 78 (26%) |
| \$20 to <\$50 thousand | 89 (30%) |
| \$50 to <\$90 thousand | 57 (19%) |
| \$90 thousand or more | 73 (25%) |
| Household Food Security<sup>c</sup> |  |
| High food security | 206 (69%) |
| Marginal food security | 42 (14%) |
| Low food security | 37 (12%) |
| Very low food security | 14 (5%) |
| Household Occupants | **Mean (SD)** |
| Number of adults | 2.0 (0.63) |
| Number of children | 2.2 (1.4) |

Percentages may not total to 100% due to rounding.
<sup>a</sup> Includes adoptive mother (n=3), grandmother (n=1), other, non-relative (n=1)
<sup>b</sup> Includes “partner and I not living together” (n=4), separated (n=5), and divorced (n=6).
<sup>c</sup> Food security was assessed using the 18-item USDA scale.
Information was missing for marital status (n=3), caregiver education (n=1), household income (n=3), and household food security (n=1)

**Figure 3:**
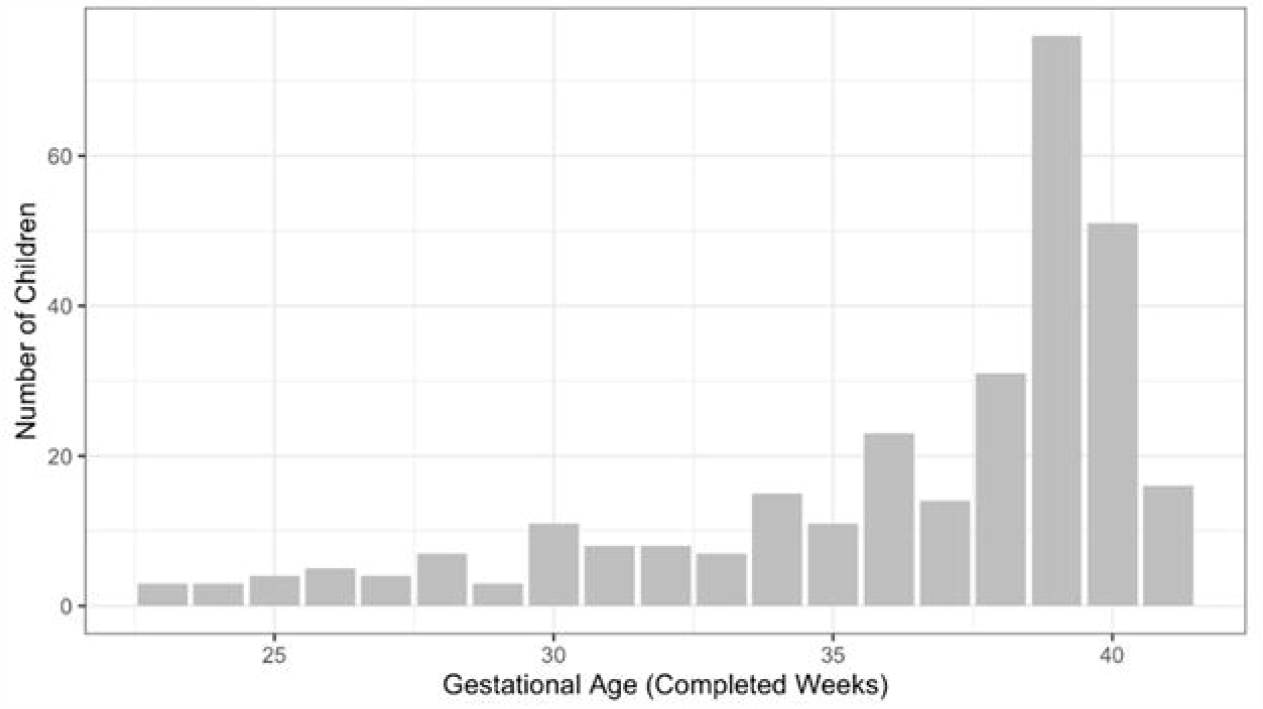
Distribution of gestational age (weeks’ completed gestation at birth) among children in the Play & Grow Cohort (n=300)

### 3.2. Child and Caregiver Weight Status

At birth, children weighed between 520 grams and 5310 grams; 21% were born at less than 2000 grams (Table 2). At enrollment, children weighed a mean (SD) of 10.9 (1.4) kg and were 79.7 (3.7) cm in length. Their BMI-for-age z-scores ranged from -2.36 to 3.47; more children (n=31) had high BMI-for-age z-scores (above 2) than children (n=2) who had low BMI-for-age z-scores (below -2). Caregiver anthropometric measurements and weight status are shown in Table 2. Almost half (48%) of caregivers had a BMI ≥ 30kg/m^2^.

**Table 2:** Child and Caregiver Anthropometric Measurements.

| **Child** | **N (%)** |
| --- | --- |
| Birthweight, grams<sup>a</sup> |  |
| <1000 | 20 (7%) |
| 1000 to <2000 | 41 (14%) |
| 2000 to <3000 | 76 (26%) |
| 3000 to <4000 | 144 (48%) |
| ≥4000 | 17 (6%) |
| Visit 1 anthropometric measurements | **Mean (SD)** |
| Length, cm | 79.7 (3.7) |
| Weight, kg | 10.9 (1.4) |
| WHO BMI-for-age z-score<sup>b</sup> | 0.75 (0.98) |
| WHO BMI-for-age z-score category<sup>b</sup> | **N (%)** |
| Underweight (BMI z-score <-2) | 2 (0.7%) |
| Healthy weight (BMI z-score -2 to <1) | 180 (60%) |
| Possible overweight (BMI z-score 1 to <2) | 86 (29%) |
| Overweight and obesity (BMI z-score ≥2) | 31 (10%) |
| **Caregiver** | **Mean (SD)** |
| Anthropometric measurements |  |
| Height (m) | 1.64 (0.072) |
| Weight (kg)<sup>c</sup> | 82.7 (23.3) |
| BMI (kg/m<sup>2</sup>)<sup>c</sup> | 30.6 (7.9) |
| Weight status<sup>c</sup> | **N (%)** |
| Underweight (BMI <18.5) | 6 (2%) |
| Healthy weight (BMI 18.5 to <25) | 79 (28%) |
| Overweight (BMI 25 to <30) | 63 (22%) |
| Obesity (BMI ≥ 30) | 139 (48%) |

N=299; excludes 1 caregiver-child dyad who did not complete visit 1. Percentages may not total to 100% due to rounding.
<sup>a</sup> Birthweight was not available for 1 child.
<sup>b</sup> Children's age was adjusted for preterm birth if children were born at <37 completed weeks' gestation. However, results were similar using unadjusted calendar age to calculate WHO BMI-for-age z-scores. Mean (SD) = 0.80 (0.97); BMI z-score <-2 (n=1), BMI z-score -2 to <1 (n=179), BMI z-score 1 to <2 (n=85), BMI z-score ≥2 (n=34).
<sup>c</sup>Excludes caregivers (n=12) who were pregnant or not measured at visit 1.

### 3.3. Comparison of Sociodemographic characteristics by Child Gestational Age

Children born preterm (<37 weeks) were similar to children born at term (≥ 37 weeks) with respect to most child and caregiver characteristics (Table 3). However, although not statistically significant (P=0.09), the sex ratio among term children includes more boys than girls (Table 3). Children born preterm were smaller as toddlers than children born at term, but the groups did not differ in distribution by weight status and the prevalence of overweight and obesity was similar in each group (Table 3). Children born preterm were more likely to live in households with annual incomes below $20,000 and less likely to live in food secure households (Table 3).

**Table 3:**
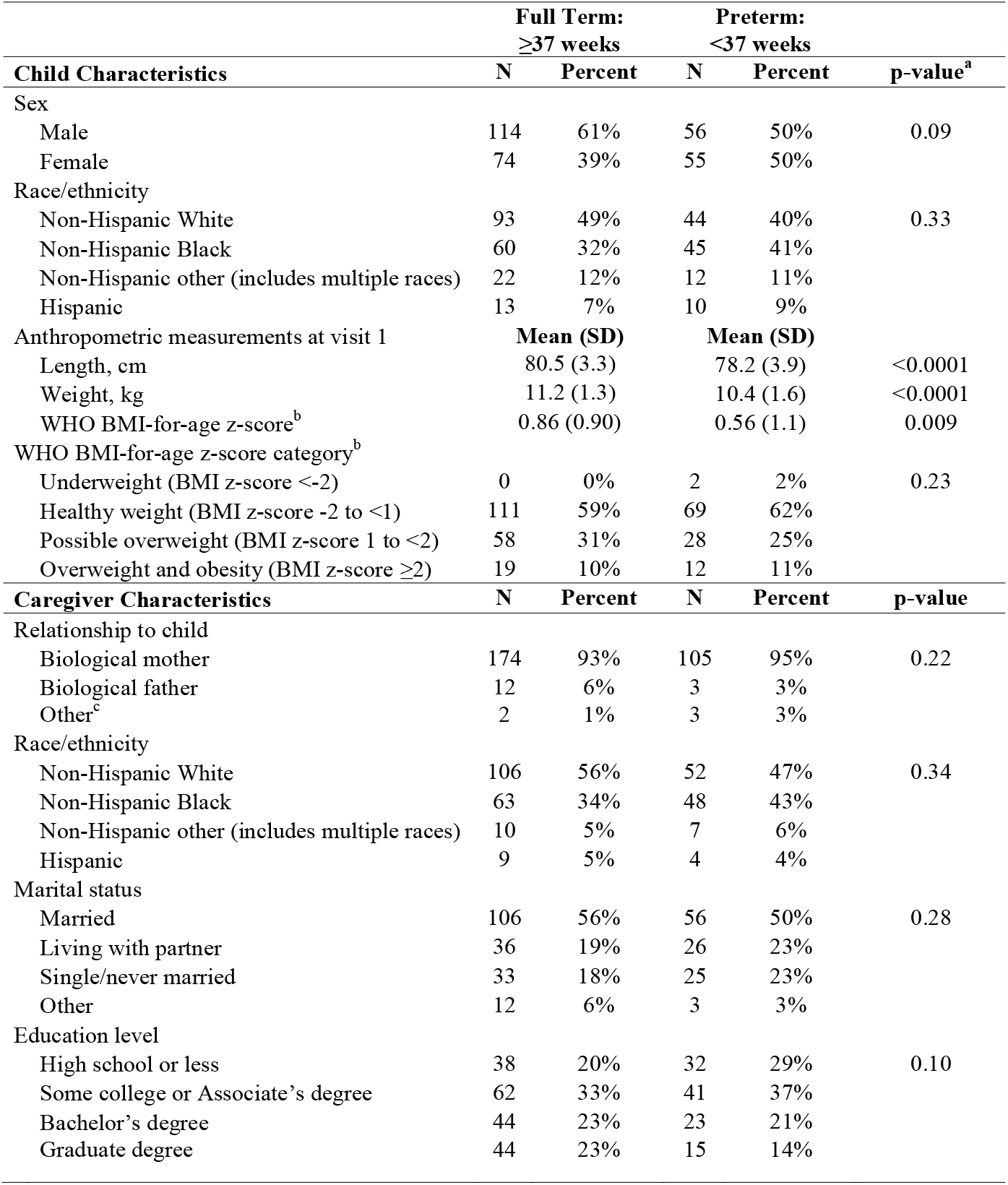
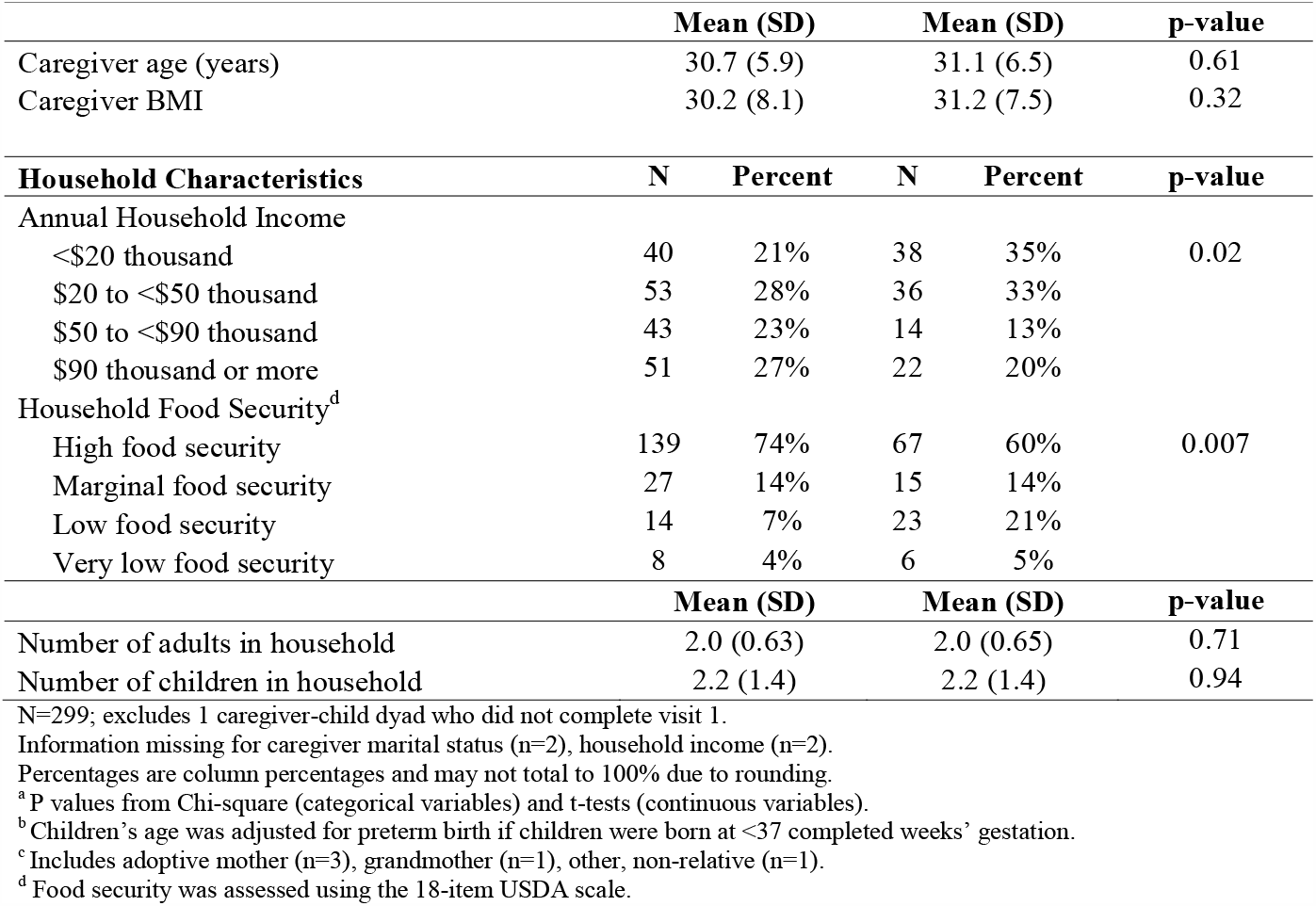
Comparison of child and caregiver characteristics by gestational age at birth.

### 3.4. Participation in Home Visit at 24 Months

Data collection for the 24-month study visit concluded in December 2019. Of the 300 caregiver-child dyads enrolled, 293 (98%) participated in the second study visit. Children were a mean (SD) calendar age of 24.0 (0.9) months. Home visits were completed with 284 families and an additional 9 families participated via online questionnaire.

## 4. DISCUSSION

The Play & Grow cohort provides an opportunity to prospectively observe caregivers and children in mealtime and non-mealtime contexts, to characterize trajectories of parent-child mealtime interactions across the toddler to preschool period relative to parent-child interactions in play, and to determine associations with self-regulation and risk for childhood obesity. The study design includes paired laboratory and home visits at each developmental period (i.e., toddler, preschool-age) to capitalize on the strengths afforded by the different assessment locations (i.e., greater standardization in laboratory vs. a more naturalistic home environment). The cohort of children and caregivers is diverse relative to gestational age at birth, race and ethnicity, parent age and education, and household income. Notably, 17% reported low or very low food security, comparable to the overall 2018 US estimate of 14% for families with young children (Coleman-Jensen, Rabbitt, Gregory, & Singh, 2019). Ohio has higher prevalence of food insecurity, obesity, and preterm birth than the United States overall (United Health Foundation, 2019). The families in the study are diverse relative to family composition, living arrangements, and home environments based on observations made in children’s homes.

Each year, more than 380,000 US children are born before 37 weeks’ completed gestation (Martin, Hamilton, Osterman, & Driscoll, 2019). Since reaching a nadir of 9.6% in 2014, the US preterm birth rate has risen during each of the last 4 years and is currently 10.0% (Martin et al., 2019). Children born preterm have risks for obesity that are similar to children born at term, and preterm children are at higher risk for deficits in self-regulation. In part because preterm infants are more excitable, irritable, and have greater difficulty directing attention and regulating arousal (Mouradian, Als, & Coster, 2000), the stresses of parenting these children are also heightened (Voigt et al., 2013).

The public health challenge of preventing obesity is now focused on the 0 to 5-year-old age range (Lumeng et al., 2015). Parent engagement is critical to the success of interventions and health promotion strategies targeting young children (Epstein, Paluch, Roemmich, & Beecher, 2007). But parents may not view their overweight child as overweight (Lundahl, Kidwell, & Nelson, 2014), or be motivated by obesity interventions that are focused on eating and activity. Further, the perceptions and expectations for children’s growth and weight status may differ for parents of preterm children because the early focus of these children’s parents and clinicians was ensuring their weight gain and survival (Estroff, Yando, Burke & Snyder, 1994). All parents may be more motivated to have a positive relationship with their child and to foster their child’s self-regulation than they are about children’s risk for obesity (Burdette & Whitaker, 2005). The Play & Grow cohort will contribute needed information about the home environments of toddlers and parenting practices that may influence children’s weight and self-regulation as they grow. We expect to find that parent-child interactions assessed during mealtimes more robustly predict obesity than parent-child interactions assessed during play, and that children’s emotional self-regulation could present a modifiable target for obesity prevention efforts. We enrolled a diverse cohort of 300 caregiver-toddler dyads and assessed >95% of families at 18 and 24 months in our laboratory and their homes. Data collection is ongoing; components of the preschool-age visits are outlined briefly below.

### 4.1. Protocol for Preschool Phase Study Visits: 36 Months and 42 Months

The preschool-phase of the study began in June 2019 when children were three years old. These study visits include video-recordings of parent-child interaction in mealtime and non-mealtime settings in the home and the lab, and direct assessment of children’s self-regulation across multiple domains. Visits are scheduled ± one week of the child’s age. The 36-month home visit included caregiver questionnaires, anthropometric measurements of the child (as described previously) and video-recording of caregiver and child playing with a standardized set of developmentally appropriate toys (barn with animals, alphabet puzzle, picture book).Trained staff completed the Home Observation for Measurement of the Environment-Short Form inventory (Caldwell & Bradley, 1984), to estimate the quality and quantity of cognitive stimulation and emotional support available to the child at home. Children’s receptive vocabulary was assessed with the Peabody Picture Vocabulary Test-Revised (Dunn, 2018) to provide a measure of early cognitive ability. As at the 24-month visit, the 36-month visit concluded with video-recording of a family meal. We are coding parent-child interaction during mealtimes at 24 and 36 months to assess stability and change in the emotional climate of family meals and better understand the range of mealtime experiences and parenting practices to which contemporary toddlers and preschool-age children are exposed. The duration of the 36-month visit was between 90 and 120 minutes.

The preschool-phase of the study concludes with a laboratory visit at 42-months. In addition to caregiver questionnaires and anthropometric measurements, trained staff administer a series of standardized executive function tasks including delay of gratification in food and non-food settings, inhibitory control, attention and emotion-regulation (Kochanska, Murray, Jacques, Koenig, & Vandegeest, 1996; Weintraub et al., 2013). The visit finishes with video-recording of a buffet-style meal designed to observe parent-child mealtime interactions in a lab setting. The meal includes a large variety and quantity of foods including hot and cold entrees, salad, beverages, snacks, and desserts which span the spectrum of nutritional quality. The caregiver and child serve themselves from the buffet and are provided 25 minutes to eat without interruption.

### 4.2. Strengths & Limitations

The Play & Grow cohort is unique in directly observing and coding parent-child interaction during mealtime and non-mealtime contexts in the home and laboratory setting during the toddler and preschool-age developmental periods. Participant recruitment was by invitation and avoided biases associated with participant self-selection. Although we were successful in enrolling a diverse cohort of caregivers and toddlers across all gestational ages, we did not meet our target for very and extremely preterm children. We think that our recruitment strategy had excellent coverage of children born very and extremely preterm residing in the Central Ohio area because NCH is the predominant NICU provider in the region. Thus, we feel confident that we attempted to recruit all age-eligible children and to enroll more children born very or extremely preterm it would have been necessary to expand our geographic area or lengthen the time for recruitment. Consequently, because our budget precluded either of these options, we increased our recruitment of children born between 32- and 36-weeks’ completed gestation; overall, 37% of children enrolled in the Play & Grow cohort were born before full term. Deficits in self-regulation for very and extremely preterm children are greater than those of children born moderately preterm whose neurocognitive functioning approaches that of term children (Clark et al., 2008). Thus, we are likely to observe less variability in self-regulation at preschool age. Other limitations include the restricted geographic area from which our cohort was identified. However, Columbus Ohio is the 14^th^-most populous city in the United States and demographically reflective of the U.S. at large (US Census Bureau, 2018). Video recording of parent-child interaction at 4 time points over the 18 to 42-month developmental period with some assessments in the home and others in the laboratory will allow for estimates of the variability of children’s experiences. But the inability to assess families in visits more closely spaced in time and the likelihood that some parents will adjust their behaviors knowing they are being recorded are additional limitations.

## 5. CONCLUSIONS

The aims of this longitudinal study, which include repeated observations of children and caregivers in their homes and in our laboratory over the toddler to preschool-age period, are to determine how the quality of parent-child interactions observed in mealtime and play settings impacts children’s growth trajectories, and to identify the aspects of child self-regulation that are associated with risk for obesity. The Play & Grow Cohort is diverse and will contribute knowledge about the dynamics of mealtime interactions over time in families with young children. Findings will inform obesity prevention efforts to help parents create and maintain routines and home environments and engage in positive relationships with their young children to foster healthy growth and development.

## Data Availability

The corresponding author takes responsibility for the data and can provide original data for review as approved by the IRB.

## Abbreviations

BMI: body mass index
IQR: interquartile range
NCH: Nationwide Children’s Hospital
NICU: neonatal intensive care unit
NIH: National Institutes of Health
SD: standard deviation
TEM: technical error of measurement
WHO: World Health Organization

## Acknowledgements

We are grateful for the participation of the families and the contributions of the research staff without whom this project would not have been possible. In particular, we acknowledge the work of Jacqueline Sullivan, Grace Pelak and Kathryn Krupsky who participated in data collection, protocol development, staff training, management and quality-control; we thank Kristin Howard for assistance with analysis and creation of tables and figures.

## Author Contributions

SEA, BZ, SK, and RA designed the study, provided oversight, and obtained funding. AP managed the recruitment and data collection and drafted the manuscript. RA and SEA conducted analyses. All authors contributed to writing and revising the of the paper and approved the final submission.

## References

Anderson, S. E., Gooze, R. A., Lemeshow, S., & Whitaker, R. C. (2012). Quality of early maternal-child relationship and risk of adolescent obesity. Pediatrics, 129(1), 132–140. doi:peds.2011-0972 [pii] 10.1542/peds.2011-0972

Anderson, S. E., & Keim, S. A. (2016). Parent-child interaction, self-regulation, and obesity prevention in early childhood. Current Obesity Reports, 5(2), 192–200. doi:10.1007/s13679-016-0208-910.1007/s13679-016-0208-9 [pii]

Baidal, J. A. W., Locks, L. M., Cheng, E. R., Blake-Lamb, T. L., Perkins, M. E., & Taveras, E. M. (2016). Risk factors for childhood obesity in the first 1,000 days: A systematic review. American Journal of Preventive Medicine, 50(6), 761–779. doi: S0749- 3797(15)00752-7 [pii] 10.1016/j.amepre.2015.11.012

Belfort, M. B., Gillman, M. W., Buka, S. L., Casey, P. H., & McCormick, M. C. (2013). Preterm infant linear growth and adiposity gain: Trade-offs for later weight status and intelligence quotient. Journal of Pediatrics, 163(6), 1564–1569 e1562. doi: S0022-3476(13)00791-9 [pii] 10.1016/j.jpeds.2013.06.032

Bergmeier, H., Paxton, S., Milgrom, J., Anderson, S., Baur, L., Hill, B., … Helen, S. (2020). Early mother-child dyadic pathways to childhood obesity risk: A conceptual model. Appetite, 144(1), 104459.

Birch, L. L., Marlin, D. W., Kramer, L., & Peyer, C. (1981). Mother-child interaction patterns and the degree of fatness in children. Journal of Nutrition Education, 13(1), 17–21.

Brown, T., Moore, T. H. M., Hooper, L., Gao, Y., Zayegh, A., Ijaz, S., … et al. (2019). Interventions for preventing obesity in children. Cochrane Database of Systematic Reviews, 7. doi:10.1002/14651858.CD001871.pub4

Brownson, R. C., Hoehner, C. M., Brennan, L. K., Cook, R. A., Elliott, M. B., & McMullen, K. M. (2004). Reliability of 2 instruments for auditing the environment for physical activity. Journal of Physical Activity and Health, 1(3), 191–208.

Bruch, H. (1973). Eating disorders: Obesity, anorexia nervosa, and the person within, Basic Books. New York.

Burdette, H. L., & Whitaker, R. C. (2005). Resurrecting free play in young children: Looking beyond fitness and fatness to attention, affiliation, and affect. Archives of Pediatrics & Adolescent Medicine, 159(1), 46–50.

Caldwell, B. M., & Bradley, R. H. (1984). Home Observation for Measurement of the Environment. Little Rock, AR: University of Arkansas at Little Rock.

Casey, P. H., Bradley, R. H., Whiteside-Mansell, L., Barrett, K., Gossett, J. M., & Simpson, P. M. (2012). Evolution of obesity in a low birth weight cohort. Journal of Perinatology, 32(2), 91–96. doi:jp201175 [pii] 10.1038/jp.2011.75

Centers for Disease Control and Prevention. (2017). National Center for Health Statistics (NCHS). National Health and Nutrition Examination Survey (NHANES): Anthropometry procedures manual. Hyattsville, MD: U.S. Department Health and Human Services, Centers for Disease Control and Prevention. Retrieved February 12, 2020 from https://wwwn.cdc.gov/nchs/data/nhanes/2017-2018/manuals/2017_Anthropometry_Procedures_Manual.pdf.

Clark, C. A. C., Woodward, L. J., Horwood, L. J., & Moor, S. (2008). Development of emotional and behavioral regulation in children born extremely preterm and very preterm: Biological and social influences. Child Development, 79(5), 1444–1462. doi:10.1111/j.1467-8624.2008.01198.x

Coleman-Jensen, A., Rabbitt, M. P., Gregory, C. A., & Singh, A. (2019). Household Food Security in the United States in 2018. Retrieved February 4, 2020 from https://www.ers.usda.gov/webdocs/publications/94849/err-270.pdf?v=963.1

Cunningham, S. A., Kramer, M. R., & Narayan, K. M. V. (2014). Incidence of childhood obesity in the United States. New England Journal of Medicine, 370(5), 403–411. doi:10.1056/NEJMoa1309753

Dunn, D. M. (2018). Peabody Picture Vocabulary Test-Fifth Edition. Retrieved January 30, 2020 from https://www.pearsonassessments.com/store/usassessments/en/Store/Professional-Assessments/Academic-Learning/Peabody-Picture-Vocabulary-Test-%7C-Fifth-Edition/p/100001984.html

Epstein, L. H., Paluch, R. A., Roemmich, J. N., & Beecher, M. D. (2007). Family-based obesity treatment, then and now: Twenty-five years of pediatric obesity treatment. Health Psychology, 26(4), 381–391. doi:2007-09406-001 [pii] 10.1037/0278-6133.26.4.381

Estroff, D. B., Yando, R., Burke, K., & Synder, D. (1994). Perceptions of preschoolers’ vulnerability by mothers who had delivered preterm. Journal of pediatric psychology, 19(6), 709–721.

Fiese, B. H., & Bost, K. K. (2016). Family ecologies and child risk for obesity: Focus on regulatory processes. Family Relations, 65(1), 94–107. doi:10.1111/fare.12170

Francis, L. A., & Susman, E. J. (2009). Self-regulation and rapid weight gain in children from age 3 to 12 Years. Archives of Pediatrics & Adolescent Medicine, 163(4), 297–302.

Franz, A. R., Pohlandt, F., Bode, H., Mihatsch, W. A., Sander, S., Kron, M., & Steinmacher, J. (2009). Intrauterine, early neonatal, and postdischarge growth and neurodevelopmental outcome at 5.4 years in extremely preterm infants after intensive neonatal nutritional support. Pediatrics, 123(1), e101–109. doi: 123/1/e101 [pii] 10.1542/peds.2008-1352

Geserick, M., Vogel, M., Gausche, R., Lipek, T., Spielau, U., Keller, E., … Körner, A. (2018). Acceleration of BMI in early childhood and risk of sustained obesity. The New England Journal of Medicine, 379(14), 1303–1312. doi:10.1056/NEJMoa1803527

Graziano, P. A., Kelleher, R., Calkins, S. D., Keane, S. P., & Brien, M. O. (2013). Predicting weight outcomes in preadolescence: The role of toddlers’ self-regulation skills and the temperament dimension of pleasure. International Journal of Obesity, 37(7), 937–942. doi:ijo2012165 [pii] 10.1038/ijo.2012.165

Hack, M., Schluchter, M., Cartar, L., Rahman, M., Cuttler, L., & Borawski, E. (2003). Growth of very low birth weight infants to age 20 years. Pediatrics, 112 (1 Pt 1), e30-38.

Hales, C. M., Fryar, C. D., Carroll, M. D., Freedman, D. S., & Ogden, C. L. (2018). Trends in obesity and severe obesity prevalence in US youth and adults by sex and age, 2007-2008 to 2015-2016. JAMA, 319(16), 1723–1725. doi:2676543 [pii] 10.1001/jama.2018.3060

Hughes, S. O., Power, T. G., O’Connor, T. M., & Fisher, J. O. (2015). Executive functioning, emotion regulation, eating self-regulation, and weight status in low-income preschool children: How do they relate? Appetite, 89, 1-9. doi:S0195-6663(15)00018-5 [pii] 10.1016/j.appet.2015.01.009

Kochanska, G., Coy, K. C., & Murray, K. T. (2001). The development of self-regulation in the first four years of life. Child Development, 72(4), 1091–1111.

Kochanska, G., Murray, K., Jacques, T. Y., Koenig, A. L., & Vandegeest, K. A. (1996). Inhibitory control in young children and its role in emerging internalization. Child Development, 67(2), 490–507.

Kong, K. L., Eiden, R. D., & Paluch, R. A. (2019). Early nonfood parent-infant interactions and development of obesity in a high-risk, diverse sample. Obesity, 27(11), 1754–1760. doi:10.1002/oby.22649

Lumeng, J. C., Taveras, E. M., Birch, L., & Yanovski, S. Z. (2015). Prevention of obesity in infancy and early childhood: A National Institutes of Health workshop. JAMA Pediatrics, 169(5), 484–490. doi:10.1001/jamapediatrics.2014.3554

Lundahl, A., Kidwell, K. M., & Nelson, T. D. (2014). Parental underestimates of child weight: A meta-analysis. Pediatrics, 133(3), e689–703. doi:peds.2013-2690 [pii] 10.1542/peds.2013-2690

Martin, J. A., Hamilton, B. E., Osterman, M. J., & Driscoll, A. K. (2019). Births: Final data for 2018. National Vital Statistics Reports, 68(13), 1–46.

Matheny Jr, A. P., Wachs, T. D., Ludwig, J. L., & Phillips, K. (1995). Bringing order out of chaos: Psychometric characteristics of the Confusion, Hubbub, and Order Scale. Journal of Applied Developmental Psychology, 16(3), 429–444.

Moding, K. J., Augustine, M. E., & Stifter, C. A. (2019). Interactive effects of parenting behavior and regulatory skills in toddlerhood on child weight outcomes. International Journal of Obesity, 43(1), 53–61. doi:10.1038/s41366-018-0162-6

Mouradian, L. E., Als, H., & Coster, W. J. (2000). Neurobehavioral functioning of healthy preterm infants of varying gestational ages. Journal of Developmental and Behavioral Pediatrics, 21(6), 408–416.

NICHD Early Child Care Research Network (1999). Child care and mother–child interaction in the first three years of life. Developmental Psychology, 35(6), 1399–1413. doi:10.1037/0012-1649.35.6.1399

Perini, T. A., Oliveira, G. L. d., Ornellas, J. d. S., & Oliveira, F. P. d. (2005). Technical error of measurement in anthropometry. Revista Brasileira de Medicina do Esporte, 11, 81–85.

Ross, C. E., & Mirowsky, J. (1999). Disorder and decay: The concept and measurement of perceived neighborhood disorder. Urban Affairs Review, 34(3), 412–432. doi:10.1177/107808749903400304

Saltzman, J. A., Fiese, B. H., Bost, K. K., & McBride, B. A. (2018). Development of appetite self-regulation: Integrating perspectives from attachment and family systems theory. Child Development Perspectives, 12(1), 51–57. doi:10.1111/cdep.12254

Schlam, T. R., Wilson, N. L., Shoda, Y., Mischel, W., & Ayduk, O. (2013). Preschoolers’ delay of gratification predicts their body mass 30 years later. Journal of Pediatrics, 162(1), 90–93. doi:S0022-3476(12)00737-8 [pii] 10.1016/j.jpeds.2012.06.049

Spill, M. K., Callahan, E. H., Shapiro, M. J., Spahn, J. M., Wong, Y. P., Benjamin-Neelon, S. E., … Casavale, K. O. (2019). Caregiver feeding practices and child weight outcomes: A systematic review. The American Journal of Clinical Nutrition, 109(Supplement_1), 990S–1002S. doi:10.1093/ajcn/nqy276

Treyvaud, K. (2014). Parent and family outcomes following very preterm or very low birth weight birth: A review. Seminars in Fetal and Neonatal Medicine, 19(2), 131–135. doi: S1744-165X(13)00100-5 [pii] 10.1016/j.siny.2013.10.008

United Health Foundation. (2019). America’s Health Rankings: Annual Report. Retrieved April 14, 2020 from https://www.americashealthrankings.org/explore/annual/measure/Obesity/state/OH

U.S. Census Bureau (2018). Quick Facts Columbus Ohio; United States. United States Census Bureau. Retrieved February 18, 2020 from https://www.census.gov/quickfacts/fact/table/columbuscityohio,US/PST045218

Voigt, B., Brandl, A., Pietz, J., Pauen, S., Kliegel, M., & Reuner, G. (2013). Negative reactivity in toddlers born prematurely: Indirect and moderated pathways considering self-regulation, neonatal distress and parenting stress. Infant Behavior and Development, 36(1), 124–138. doi: S0163-6383(12)00132-4 [pii] 10.1016/j.infbeh.2012.11.002

Weintraub, S., Dikmen, S. S., Heaton, R. K., Tulsky, D. S., Zelazo, P. D., Bauer, P. J., … Wallner-Allen, K. (2013). Cognition assessment using the NIH Toolbox. Neurology, 80 (11 Supplement 3), S54–S64.

Wendland, B. E., Atkinson, L., Steiner, M., Fleming, A. S., Pencharz, P., Moss, E., … Levitan, R. D. (2014). Low maternal sensitivity at 6 months of age predicts higher BMI in 48 month old girls but not boys. Appetite, 82, 97–102. doi: S0195-6663(14)00369-9 [pii] 10.1016/j.appet.2014.07.012

WHO Multicentre Growth Reference Study Group. (2006). WHO Child Growth Standards: Length/height-for-age, weight-for-age, weight-for-length, weight-for-height and body mass index-for-age: Methods and development. Retrieved February 6, 2020 from: https://www.who.int/childgrowth/standards/technical_report/en/

World Health Organization (2020). WHO Antro Survey Analyser and Other Tools. Retrieved February 6, 2020 from https://www.who.int/childgrowth/software/en/

Wu, T. J., Dixon, W. E., Dalton, W. T., Tudiver, F., & Liu, X. F. (2011). Joint effects of child temperament and maternal sensitivity on the development of childhood obesity. Maternal and Child Health Journal, 15(4), 469–477. doi:DOI 10.1007/s10995-010-0601-z

Zheng, M., Lamb, K. E., Grimes, C., Laws, R., Bolton, K., Ong, K. K., & Campbell, K. (2018). Rapid weight gain during infancy and subsequent adiposity: A systematic review and meta-analysis of evidence. Obesity Reviews, 19(3), 321–332. doi:10.1111/obr.12632

Zvara, B. J., Mills-Koonce, W. R., Garrett-Peters, P., Wagner, N. J., Vernon-Feagans, L., & Cox, M. (2014). The mediating role of parenting in the associations between household chaos and children’s representations of family dysfunction. Attachment and Human Development, 16(6), 633–655. doi:10.1080/14616734.2014.966124

